# Global Pan-cancer serum miRNA classifier across 13 cancer types: Analysis of 46,349 clinical samples

**DOI:** 10.1101/2025.02.07.25321821

**Authors:** Pandikannan Krishnamoorthy, Madhavan Parthasarathy, Nilanjana Das, Athira S Raj, Ashok Kumar, Vikas Gupta, Saikat Das, Himanshu Kumar

**Author notes:** Corresponding author: H Kumar, Department of Biological Sciences, Laboratory of Immunology and Infectious Disease Biology, Indian Institute of Science Education and Research (IISER) Bhopal, AB-3, Room No. 220, Bhopal By-pass Road, Bhauri, Bhopal 462066, MP, India.

## Abstract

Liquid biopsy offers the minimally-invasive way of early cancer diagnosis. MicroRNAs (miRNAs) are small non-coding RNAs that show promising diagnostic potential due to their stability and their dysregulation upon different physiological conditions. However, existing cancer classifiers often rely on cohort-based comparisons, limiting their clinical utility. Extensive analyses in this study present a pan-cancer miRNA-based single-sample classifier, trained on 16,190 samples, tested across 9 independent datasets, and further validated on 8 distinct disease cohorts. The classifier leverages miRNA expression signatures to classify cancer and non_cancer samples including healthy, other diseases with high sensitivity and specificity, enabling personalized predictions. The classifier identifies cancer by evaluating the relative expression patterns of specific miRNAs, capturing neoplasm-specific dysregulation patterns independent of cohort effects. This study highlights the potential of miRNAs in robust cancer classification, offering a minimally invasive, scalable, and clinically adaptable miRNA serum resource for early cancer detection across diverse populations and malignancies.

## Main

Globally cancer remains a major health burden with nearly 20 million new cases and causing 9.7 million recorded deaths worldwide^1^. Nearly one in every six deaths recorded globally are caused by cancer and the new cancer incidences are predicted to increase up to 35 million by 2025^1,2^. However, overall cancer mortality has decreased to nearly 33% since 1991, due to the advancement in the early diagnosis, treatment methods, population-level screening^3,4^. Therefore, early diagnosis of cancer is crucial.

There are major challenges in the existing early diagnosis methods as they are invasive and cancer tissue-specific. Early diagnosis methods utilizing minimally-invasive liquid biopsy methods involving the clinical samples such as plasma, serum, and non-invasive liquid biopsy method using patient urine and saliva can play an important role in risk stratification. Majority of existing liquid biopsy methods utilize the cancer specific mutations, methylation patterns that have its own limitations such as prevalence of false-positive results due to the presence of other diseases^5^.

The MicroRNAs (miRNAs) are small non-coding RNAs that are readily present in the extracellular fluids and show differential patterns associated with clinical symptoms and with detectable tumors. ^6,7^. The miRNAs are highly stable in the blood serum and possess diagnostic potential. Several studies utilized advanced statistical methods, machine learning algorithms to identify miRNA-based classifiers in blood components such as serum and plasma. However, the biggest drawback limiting their clinical applications is that miRNAs are dysregulated in several diseases and the cancer-specific signature often overlaps with any other diseases associated with inflammation. The existing databases such as The Cancer Genome Atlas (TCGA) only consist of cancer tissue-specific omics datasets and the cancer miRNA profiles derived from the serum samples are not harmonized in a single database. There are very limited resources for the liquid biopsy transcriptome in larger sample sizes to infer any models with high confidence.

By employing the rules-based binary classification algorithms, this study identified the rules-based classifier model utilizing the set of four rules, discriminate between cancer and non_cancer samples including healthy individuals and patients from other non-infectious diseases with highest accuracy. The accuracy of the model was also validated in the independent datasets comprising the cancer and other diseases.

## Results

### Mining of Serum miRNA datasets across cancer types

TCGA lacks the miRNA expression datasets derived from blood serum and hence the public transcriptome datasets such as GEO (Gene Expression Omnibus), Array Express were queried for the eligible miRNA expression datasets derived from serum. A total of 46349 samples from the independent datasets were identified and screened for the analysis (**Supp. Fig. 1**). 490 samples were removed due to the presence of duplicates across datasets and improper labelling. The distribution of samples across each cancer type in each dataset were visualized in (**Figure 1A).** The dataset GSE211692 was utilized as the training dataset due to its higher sample size of 16190, out of which 9921 samples belong to the “cancer” class and 6269 samples belong to “non_cancer” class ^8^. The datasets GSE122497^9^, GSE106817^10,11^, GSE137140^12^, GSE113740^13^, GSE112264^14^, GSE139031^15^, GSE113486^16^, GSE164714^17^ and GSE73002^18^ were utilized as test datasets that comprise of both “cancer” and “non_cancer” class for which the distribution is visualized in (**Fig. 1A)** (**Supp. Table 1**). The datasets GSE110317^19^, GSE110651^20^, GSE134108^21^, GSE85589^22^, GSE117064^23^, GSE120584^24^, GSE150693^25^and GSE140249^26^ were considered validation datasets that comprises of either “cancer” or “non_cancer” class. Even if both classes were present, at least one class had very less sample size. The “non_cancer” class consist of diverse non-cancer diseases such as Autoimmune Hepatitis, Alzheimer’s disease (AD), Vascular Dementia (VD), Mild cognitive Impairment (MCI). The purpose of validation datasets is to test whether the classifier is able to correctly predict “non_cancer” samples with identified rules. Overall, 46349 serum samples profiled for miRNA expression were included in the study. Out of which 16190 samples, 25082 samples and 5077 samples were utilized as training dataset, test datasets and validation datasets, respectively.

**Fig. 1:**
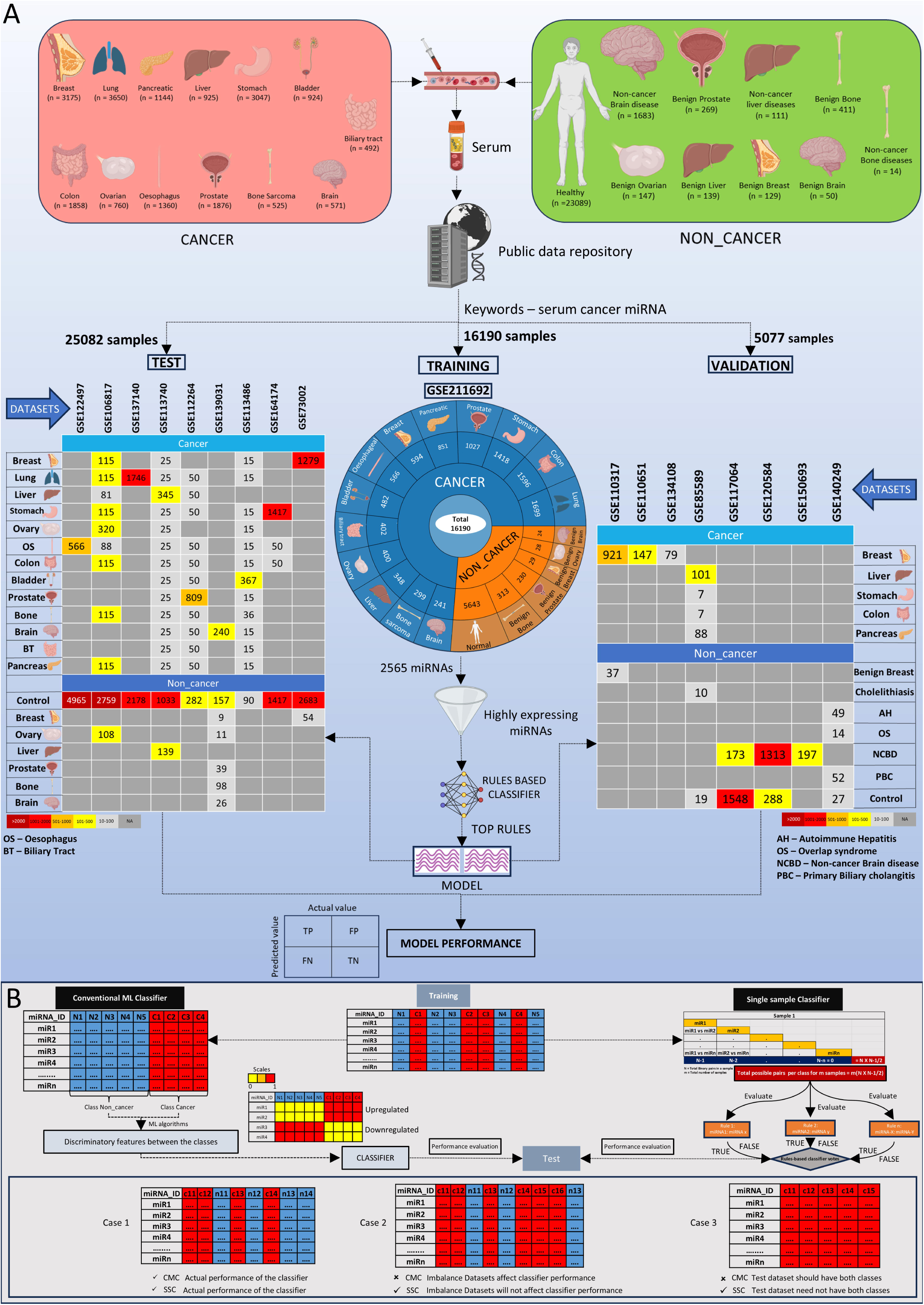
Schematic Representation of Selection of Serum miRNA datasets, samples distribution and Rules-Based single sample Classifier Model Construction. (A) Overview of serum samples distribution of “cancer” and “non_cancer.” The “cancer” samples consist of 13 cancer types and the “non_cancer” samples consist of healthy and other disease as depicted in A. The number in parentheses represent number of samples. Overall 46,349 samples were included that were split into, Training data (16,190 samples) to build the rules-based machine learning classifier model and the resulting model is tested in, Test datasets (25,082 samples) obtained from 9 independent datasets contain both cancer and non_cancer classs. The robustness of the model as single sample classifier in identifying even cancer and non_cancer samples separately is validated in (iii) 8 independent Validation datasets (5077 samples) that comprise either “cancer” or “non_cancer” that includes autoimmune diseases, neurological disorders and cerebrovascular disorders. (B) Superiority of the single-sample classifier (SSC) approach over the conventional machine learning (ML) classifier (CMC) is depicted in three cases of equal sample size between the classs (Case 1), class imbalance (Case 2) and the need of equal number of classes (Case 3).

The conventional machine learning classifier analyzes overall miRNA (features) expression variation across the samples that belong to different class(es) and identifies discriminatory features through statistical or machine learning algorithms. The top discriminatory features, comprising significantly dysregulation between each class, will be utilized to build the classifier model. The performance evaluation of this classifier can be affected by class imbalance, batch effects between independent datasets, requirement of appropriate normalization techniques and more importantly test and validation datasets require the same number of classes as in training datasets. Rules-based single sample classifier utilized in this study identifies pairwise comparisons of each miRNA with other miRNAs and establishes the relationship within a single sample. The rules-based classifier in the format of expression of miRNA A greater than miRNA B denotes “cancer” class and reverse is “non_cancer” class. The rules-based classifier obtained through this approach can be validated in independent datasets and their performance will not be affected by batch effects or normalization methods and also the validation datasets need not have the same number of classes. For clinical diagnosis and screening in a large population, this approach can be valuable. The differences between our approach and the conventional approach is depicted in (**Fig. 1B).**

### Top Serum miRNA rules that can discriminate between “cancer” and “non_cancer” class

The training dataset GSE211692 comprises samples belonging to 13 different cancer types and non_cancer samples along with benign samples. Distribution of cancer types in the dataset is depicted through Lollipop plot (**Fig. 2A**). The expression of 2565 miRNAs across 16190 samples were subjected to filtering in which miRNAs with minimal expression in more than 10% of the samples were discarded. 211 miRNAs remained for the further analysis (**Fig. 2B**). The top scoring pairs (TSP) were identified for each sample per class and the top rules identified through each class were identified. The segregation between the “cancer” and the “non_cancer” class was visualized through the t-SNE plot (**Fig. 2C)**. A total of 4 TSPs were identified to segregate the “cancer” and “non_cancer” classes based on the scores that depicts how closely the pairs are associated with the class (**Fig. 2D**). miR-4258 > miR-4730, miR-663a > miR1228-5p, miR-6787-5p > miR-6800-5p and miR-1469 > miR-1268a are the top rules that are specific for “cancer” class and confusion matrices denotes the relationship between actual and predicted labels for the rules (**Fig. 2E – 2H**). Overall, the classification is best for the first two rules – miR-4258 > miR-4730 and miR-663a > miR-1228-5p, followed by other two rules of the training data.

**Fig. 2:**
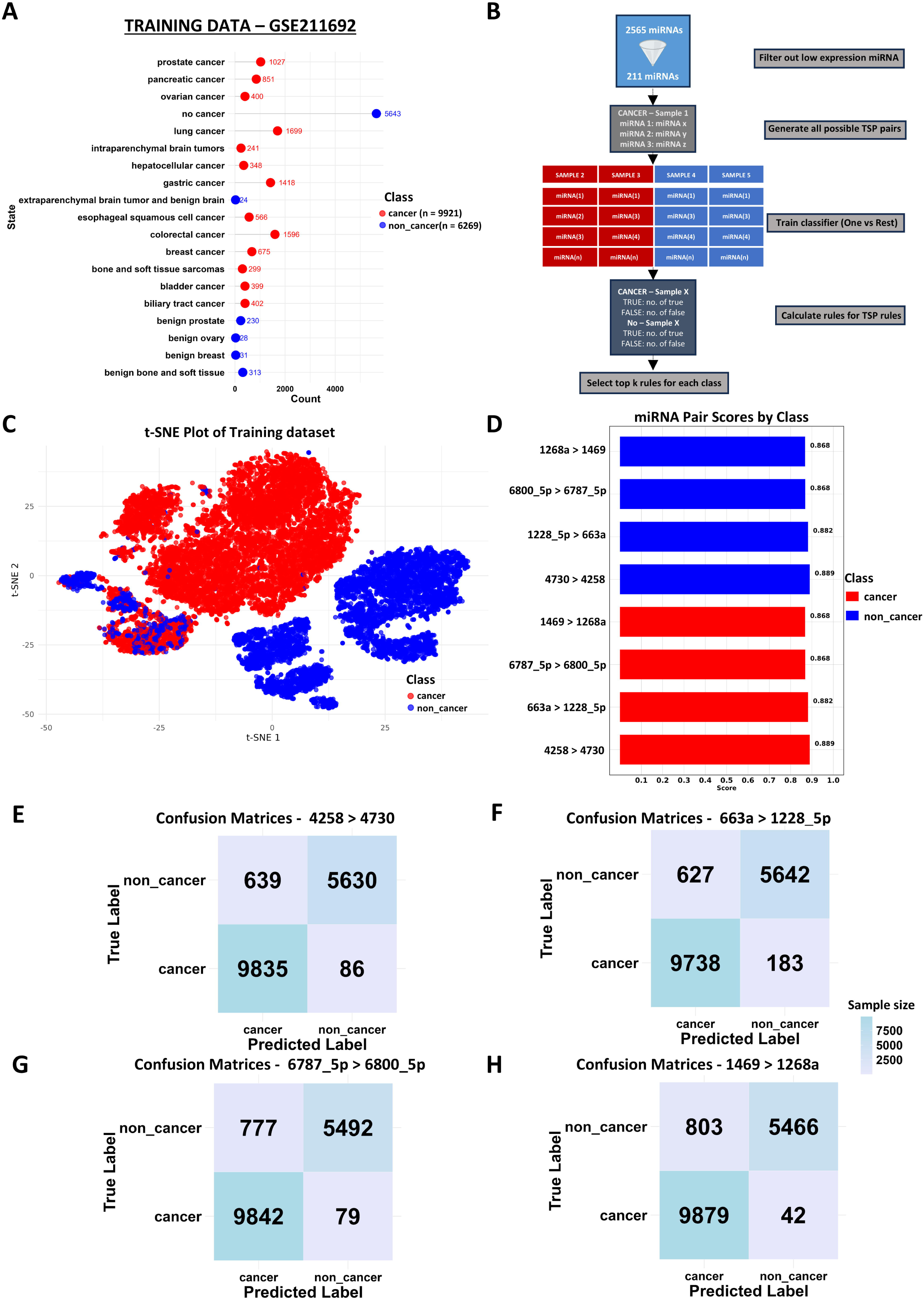
Identification of top miRNAs rules combination to classify “cancer” and “non_cancer” serum samples. (A) Lollipop plot illustrates the distribution of different types of “cancer” (red) and “non_cancer” (blue) types present in the training data GSE211692. (B) Rules-based training approach is illustrated in the schematic. miRNA expression processing phase comprises filtering, generation of top-scoring pairs (TSPs), training process of classifiers using one versus rest scheme and calculation of scores of each TSP pair. The top k rules for each class were identified. (C) The segregation of the “cancer” (red) and “non_cancer” (blue) classes are illustrated through the T-distributed Stochastic Neighbor Embedding (t-SNE) plot. (D) The Final four score rules for each class identified were denoted and their comparison rules in “cancer” and “non_cancer” classes are denoted. (E) The confusion matrices denote the relation between the predicted and the actual classes for rules: (E) 4258 > 4730, (F) 663a > 1228_5p, (G) 6787_5p > 4800_5p and (H) 1469 > 1268a.

### Construction of Serum pan-cancer single sample classifier using four rules

The expression of the eight miRNAs forming four rules were visualized through beeswarm plot (**Fig. 3A**). There is a clear distinction between the expression pattern of the miRNA pairs in “cancer” and “non_cancer” class. Notably, expression level of each miRNAs is higher and hence it has higher diagnostic potential. The overall performance metrics of each of the rules are visualized using heatmap (**Fig. 3B**). The rules 4730>4258 and 663a>1228_5p performs better than the other two rules individually. Relying on any one rules can affect the specificity across diverse disease conditions and hence the classifier model was built based on the vote-based approach where more than two rules have to be satisfied to be predicted as “cancer”, and predicted as “non_cancer” when two or less than two rules are satisfied (**Fig. 3C**). The pan-cancer classifier with this voting-based approach was evaluated in the training dataset and the classifier performance improved as a whole as depicted in the confusion matrix (**Fig. 3D**). The confusion matrix depicts that 9897 samples out of 9921 samples were correctly predicted as “cancer”, whereas 5626 samples out of 6269 samples were correctly predicted as “non_cancer”. Only 24 “cancer” samples were wrongly predicted as “non_cancer”, whereas 643 “non_cancer” samples were predicted as “cancer”. The heatmap denotes correct predictions (light green) and incorrect predictions (light red) across 16190 samples of training data (**Fig. 3E**) (**Supp. Table 2**). The cancer types among the cancer class are also highlighted. Overall, the 4-rules based serum pan-cancer classifier predicts the “cancer” and “non_cancer” with higher accuracy while the performance of classifier is affected while predicting certain “non_cancer” samples that are predicted as “cancer”.

**Fig. 3:**
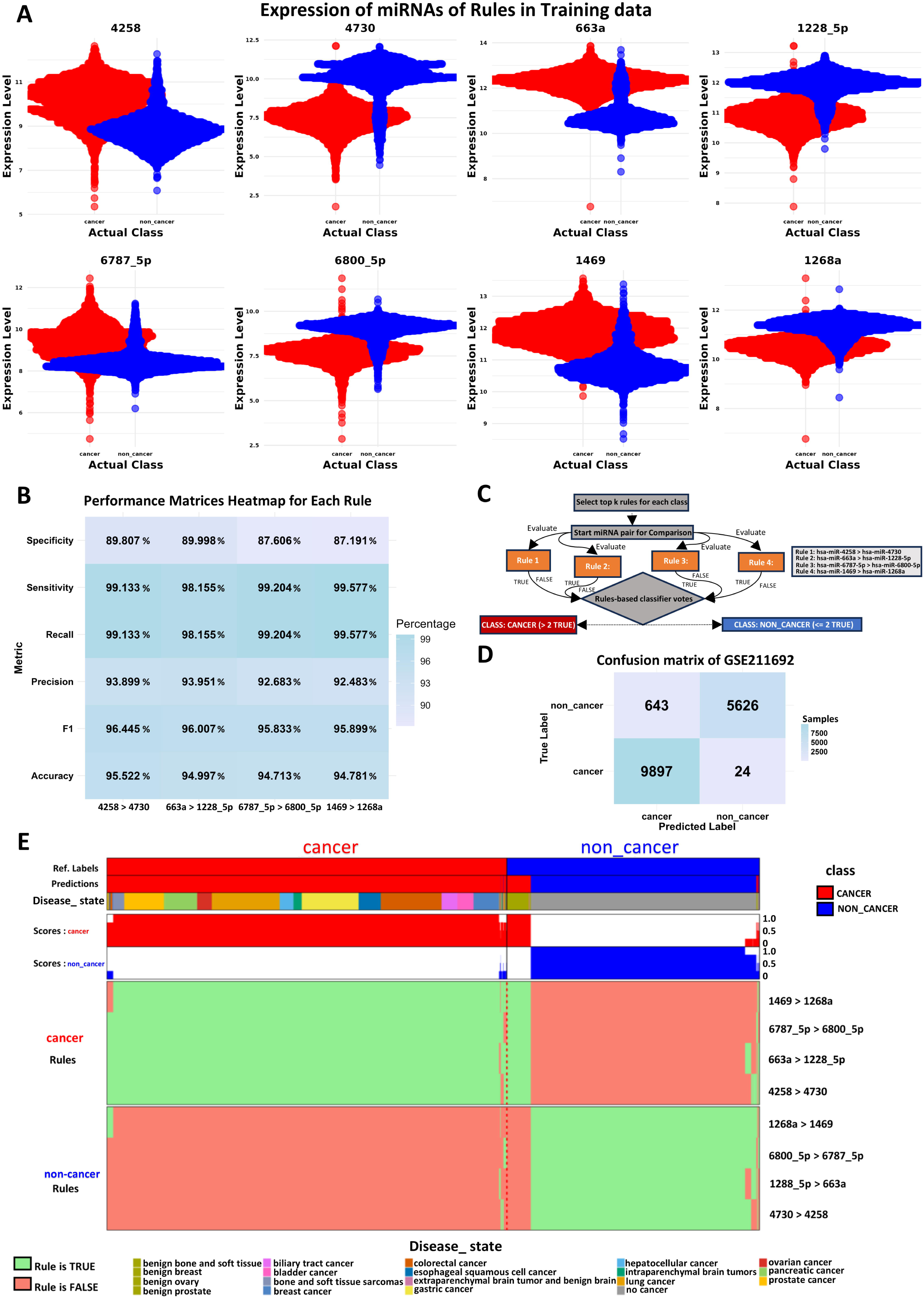
Construction of pan-cancer serum miRNA rules-based classifier. (A) The expression of eight miRNAs that form the four rules across the “cancer” and “non_cancer” classes were plotted through the bee-swarm plots where red denotes “cancer” and blue denotes “non_cancer” class. (B) The individual rules and their classification metrics such as Specificity, Sensitivity, Recall, Precision, F1 value and accuracy were plotted as heatmap. (C) Schematic of the classifier construction incorporating the voting-based classification in which the samples are predicted as cancer if more than two rules are satisfied and “non_cancer” if less than 2 or equal to 2 satisfied rules. (D) Confusion matrix denotes the relationship between the predicted and true classes when all four rules are combined as a classifier. (E) Heatmap denotes the classification performance of all the four rules that are selected as classifier. Prediction Scores for each sample were plotted in red and green for “cancer” and “non_cancer” respectively. The distribution of the disease types and the corresponding predictions were depicted at the bottom of the heatmap.

### Classifier performance evaluation in independent test datasets

The performance of the classifier is tested in 9 independent test datasets. Test datasets comprise of both “cancer” and “non_cancer” classes samples derived from individual studies that are not part of the training dataset. The overall distribution of disease state of both the “cancer” and “non_cancer” class of samples across all the 9 datasets were visualized through the tile plot (**Fig. 4A**). The distribution of samples between “cancer” and “non_cancer” class were visualized through t-SNE plots for all the 9 test datasets (**Supp. Fig. 2 A-I**). The expression levels of eight miRNAs that are the part of pan-cancer classifier was plotted and visualized through beeswarm plots for all the test datasets that clearly tells these miRNAs are having higher expression and also the change in the expression patterns between two classes were distinct (**Supp. Fig. 3 A-I**). The classification of individual rules was plotted through the confusion matrices for all test datasets (**Supp. Fig. 4 B-J**). Individual rules experience varied classification results. The status of predictions by the pan-cancer classifier model was visualized through the confusion matrices (**Fig. 4B**). As observed in the training datasets, the classifier performance is good in predicting “cancer” samples as “cancer” accurately in almost all the datasets. The performance of the classifier was poor while predicting few “non_cancer” samples as the classifier misclassify them as “cancer”. The error is more in GSE112264 dataset as 242 samples out of 262 “non_cancer” samples were predicted as “cancer”. However, all the 1309 cancer samples were predicted as “cancer”. The heatmap denotes the prediction results of all the 25082 samples present in nine test datasets (**Fig. 4C**) and details of the predictions of all samples are provided in (**Supp. Table 3**). The disease state distribution for the “cancer” and “non_cancer” class were also observed through this plot. The heatmap of performance metrices of the pan-cancer classifier over all test datasets shows poor performance of the GSE112264 dataset in terms of F1 score, Recall and sensitivity due to misclassification of the “non_cancer” samples as “cancer” samples (**Fig. 4D**). The diagnostic performance of the pan-cancer classifier was visualized through area under the Receiver Operating curve (ROC) plots and that shows poor performance in GSE112264 dataset (**Fig. 4E**). Overall, pan-cancer classifier predicts “cancer” samples as “cancer” accurately in 9 independent datasets whereas the performance was poor in prediction of “non_cancer” samples.

**Fig. 4:**
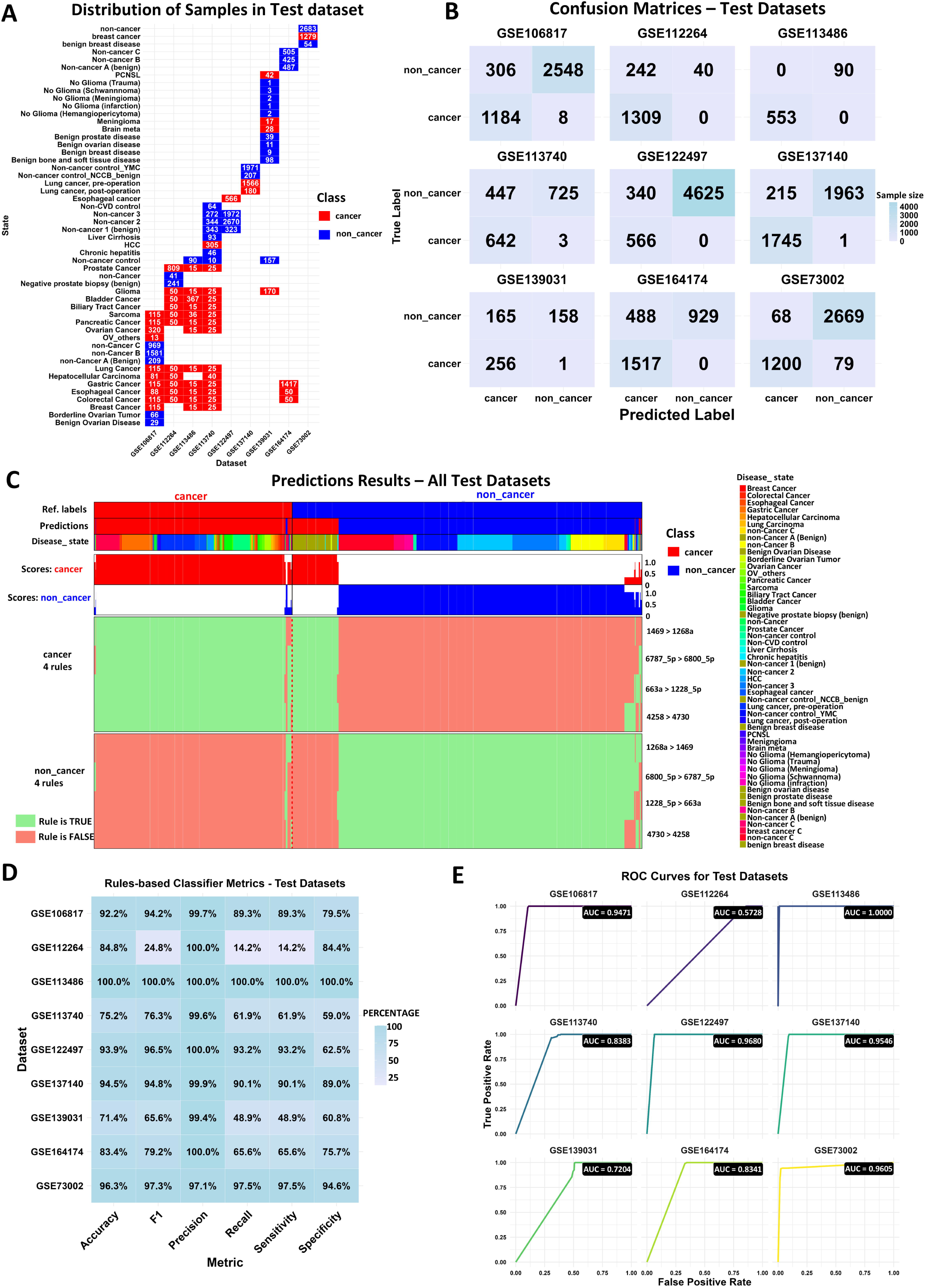
Evaluation of pan-cancer classifier performance in independent test datasets. (A) Tiles plot denotes the distribution and all the types of samples present across all the 9 test datasets and the tile color represents the class-red for “cancer” and blue for “non_cancer”. (B) The performance of the classifier in 9 individual datasets were plotted as confusion matrices that denote the relationship between the predictions (x-axis) and the true label (y-axis). (C) The heatmap plot depicts the prediction results of all the 25082 samples across 9 independent test datasets comprising different disease states and prediction Scores for each sample were plotted in orange and light green for “cancer” and “non_cancer” respectively. (D) The classifier performance in individual datasets and their classification metrices such as Specificity, Sensitivity, Recall, Precision, F1 value and accuracy were plotted as heatmap. (E) The Receiver operator curves (ROC) to evaluate the diagnostic potential by comparing the true positive rate and false positive rate for all the dataset and the area under curve values for each of the datasets were illustrated.

### Classifier performance in validation datasets comprising other diseases

The biggest advantage of single sample classifier is that the performance of the classifier can be evaluated even in datasets without requirement of all classes present in the training process. Eight independent validation datasets were identified that has either one of the “cancer” and “non_cancer” classes or imbalanced sample sizes between both the classes. The distribution of the disease state of all the samples across the independent validation datasets were visualized through the tiles plot (**Fig. 5A**). Since the classifier performance was not up to mark in prediction of “non_cancer” class, validation datasets contain the majority of samples from other disease conditions apart the cancer. Some of the other diseases includes Alzheimer’s disease, Autoimmune diseases, Dementia, Mild cognitive impairment, Auto-immune Hepatitis. The predictions of each rules were identified through the confusion matrices (**Supp. Fig. 5 B-I**). The performance of the pan-cancer classifier in the validation datasets was visualized through the confusion matrices (**Fig. 5B**). The dataset GSE110317 comprises of 958 samples, out of which 921 were breast cancer and 37 were benign breast cancer samples. All the 37 benign breast cancer samples were predicted as “cancer” instead of “non_cancer” class. However, 918 out of 921 breast cancer samples were predicted as “cancer”. GSE110651 datasets comprises of 147 metastatic breast cancer samples and 145 of them were correctly predicted as “cancer”. GSE117064 dataset comprises of 1785 cerebrovascular diseases and non-CVD control samples and 1776 samples were correctly predicted as “non_cancer” class. GSE120584 comprises of 1601 samples of neurological disorders such as Alzheimer’s disease, Dementia and non-disease controls. 1596 samples were correctly predicted as “non_cancer” class by the classifier. GSE134108 dataset comprises of 71 samples of both the metastatic and non-metastatic brain cancer. All of the samples were perfectly predicted as “cancer”. GSE140249 dataset comprises of 142 samples of autoimmune hepatitis, Biliary Cholangitis and 133 of these samples are predicted as “non_cancer” class. GSE150693 dataset comprises of 197 Mild cognitive Impairment patients with or without Alzheimer’s disease conversion and 196 of them were predicted accurately as “non_cancer” class. The GSE85589 dataset comprises of 232 samples of both “cancer” and “non_cancer” samples that includes pancreatic cancer, Cholangiocarcinoma, colorectal cancer and non-cancer controls. 192 of 203 cancer samples were predicted correctly as “cancer”. However, all the 29 non-cancer samples were predicted as “cancer”. The overall prediction results of validation datasets and the disease state distribution was observed through the heatmap (**Supp. Fig. 5J**) and details of the prediction results of each sample is provided in (**Supp. Table 4**). The number of correct (light green) and incorrect predictions (red) in each of validation datasets were visualized through bar plot (**Fig. 5C**). The distribution of disease state for only the incorrect predictions was visualized through the tiles plot (**Fig. 5D**).

**Fig. 5:**
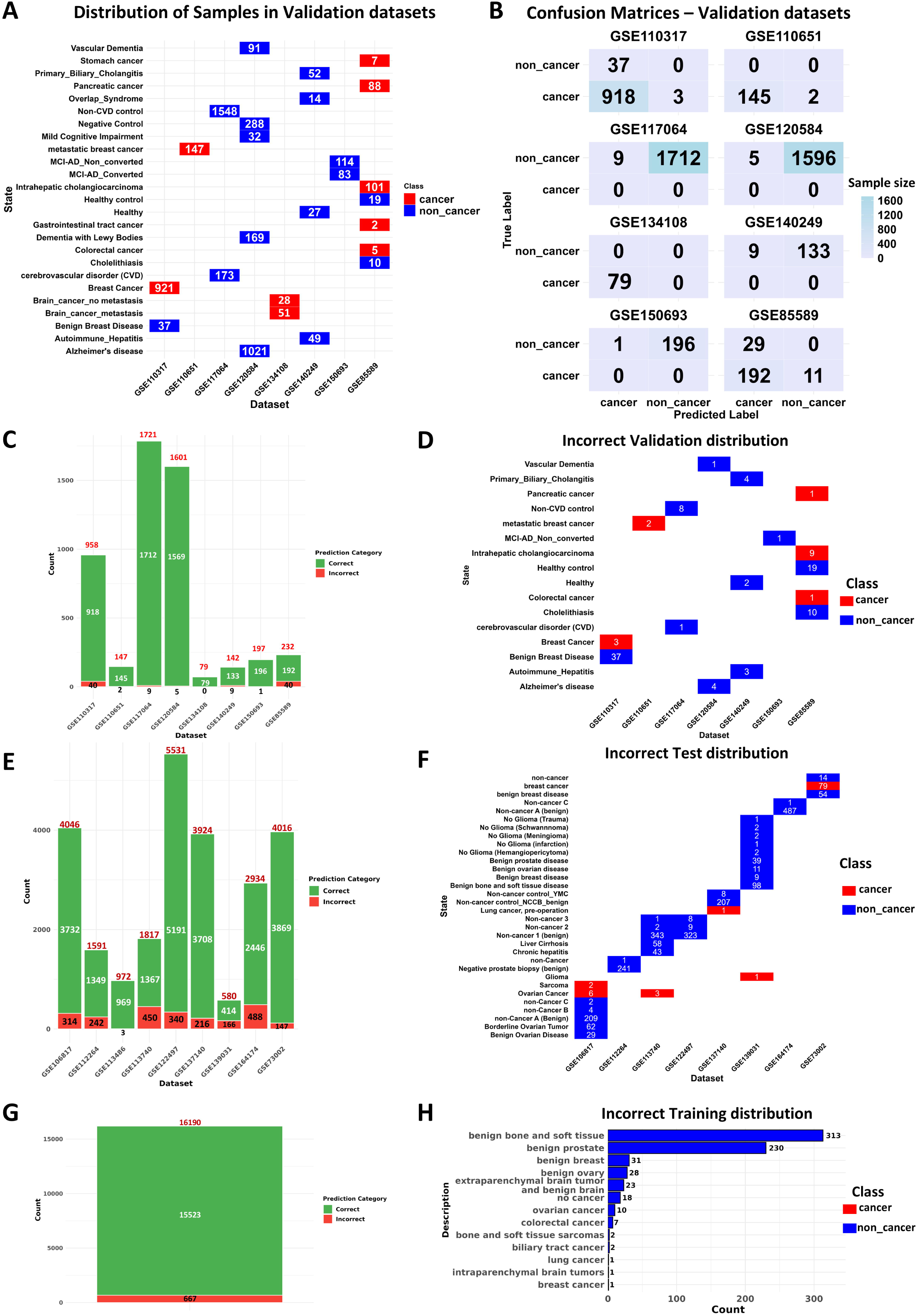
Pan-cancer classifier robustness as single-sample classifier and its influence on benign samples: (A) Tiles plot denotes the distribution and the number of all types of samples present across all the 8 validation datasets and the tile color represents the class-red for “cancer” and blue for “non_cancer.” (B) Evaluation of pan-cancer classifier performance in individual datasets were plotted as confusion matrices that denote the relationship between the predicted and true labels (C) The Bar plot denotes the number of samples predicted correct and incorrect by the classifier in the validation datasets where the light green represents the correct predictions and orange represents the incorrect prediction. The values on each bar represent the total number of samples in each dataset. (D) The tile plot depicts the distribution of sample types that are predicted incorrectly in the validation datasets. All the benign samples in the GSE110317 are predicted as cancer. (E) The Bar plot denotes the number of samples predicted correct and incorrect by the classifier in the test datasets where the light green represents the correct predictions and orange represents the incorrect prediction. The values on each bar represent the total number of samples in each dataset. (F) The tile plot depicts the distribution of sample types that are predicted incorrectly in the test datasets. (G) The Bar plot denotes the number of samples predicted correct and incorrect by the classifier in the training data where the light green represents the correct predictions and orange represents the incorrect prediction. The values on each bar represent the total number of samples in each dataset. (H) The bar plot depicts the distribution of sample types that are predicted incorrectly in the training dataset.

Interestingly, the pan-cancer classifier wrongly predicted as cancer in 37 non-cancer samples and all of them were benign breast cancer samples in the dataset GSE110317. All the incorrect predictions of the classifier in test datasets and training datasets were explored to see if there is any pattern among the wrong predictions. The number of incorrect predictions in the test and training datasets were visualized through bar plots (**Fig. 5E)** and (**Fig. 5G**) respectively. The distribution of the disease state of the incorrectly predicted samples in test datasets and training dataset were visualized through the tile plot (**Fig. 5F**) and bar plot (**Fig. 5H**) respectively. In the test datasets, 314 wrong predictions were observed in GSE106817, out of which 306 samples were of benign origin. 29 benign ovary, 62 borderline ovary and 215 benign non-cancer samples were predicted as “cancer”. Similarly, all 242 wrong predictions of GSE112264 were identified as benign samples being predicted as “cancer”. GSE113486 dataset has no benign samples and there are only 3 wrong predictions. GSE113740 dataset has 450 wrong predictions, out of which 343 samples were benign in nature and 101 samples were liver cirrhosis, chronic hepatitis patients. 340 samples predictions were wrong in the dataset GSE122497, out of which 323 samples were of benign nature. 215 of 216 wrongly predicted samples of GSE137140 dataset were benign class. 157 out of 166, 487 out of 488 samples wrong predictions, in GSE164174 and GSE164174 datasets respectively were from benign samples. GSE73002 predicted 79 breast cancer samples as “non_cancer”. In training dataset also, 625 out of 643 wrongly predicted samples were benign samples (**Fig. 5H**). Overall, the benign samples were segregated to “cancer” rather than “non_cancer” samples by the pan-cancer classifier (**Fig. 3E**, **Fig. 4C, Supp. Fig. 5J**). Healthy control samples or samples from other diseases were predicted correctly as “non_cancer” class, as observed in the validation datasets.

### Classifier performance increases when the benign samples were assigned in “cancer” class

Since benign samples affect the performance of the pan-cancer classifier, the distribution of “cancer”, “non_cancer” and “benign” classes in the test datasets were observed through the t-SNE plots (**Supp. Fig. 6 A-J)**. It was observed that “benign” samples indeed form a distinct distribution that are close to cancer samples in many datasets. The classifier performance over test datasets were evaluated after assigning the “cancer” class to “cancer/benign” class that comprises of cancer and benign samples. The “non_cancer” class was restructured to “non_cancer/non_benign” samples that lacks the benign samples. The individual rules performances were visualized through the confusion matrices and it is observed that the performance of all the four rules improved when “benign” is included in the “cancer” class (**Supp. Fig.7**). The pan-cancer classifier performance increased in identifying the “cancer/benign” class in comparison with the performance when benign is included in the “non_cancer” class (**Fig. 6A, 6B)** (**Supp. Table 5**). The total number of samples in the training datasets and the prediction results when the benign is included in “non_cancer” class (**Fig. 6C**) and when benign is included in “cancer/benign” class (**Fig. 6D**) suggest the training data classification also improved. The total number of test datasets classification results when benign samples are included in “non_cancer” class (**Fig. 6E**) and when benign samples are included in “cancer/benign” class (**Fig. 6F**) (**Supp. Table 6**) suggest overall improvement in classification results of pan-cancer classifier. The performance metrices improved for individual rules in test datasets when benign was considered in “cancer/benign” class, in comparison with the “non_cancer” class (**Supp. Fig. 8**).

**Fig. 6:**
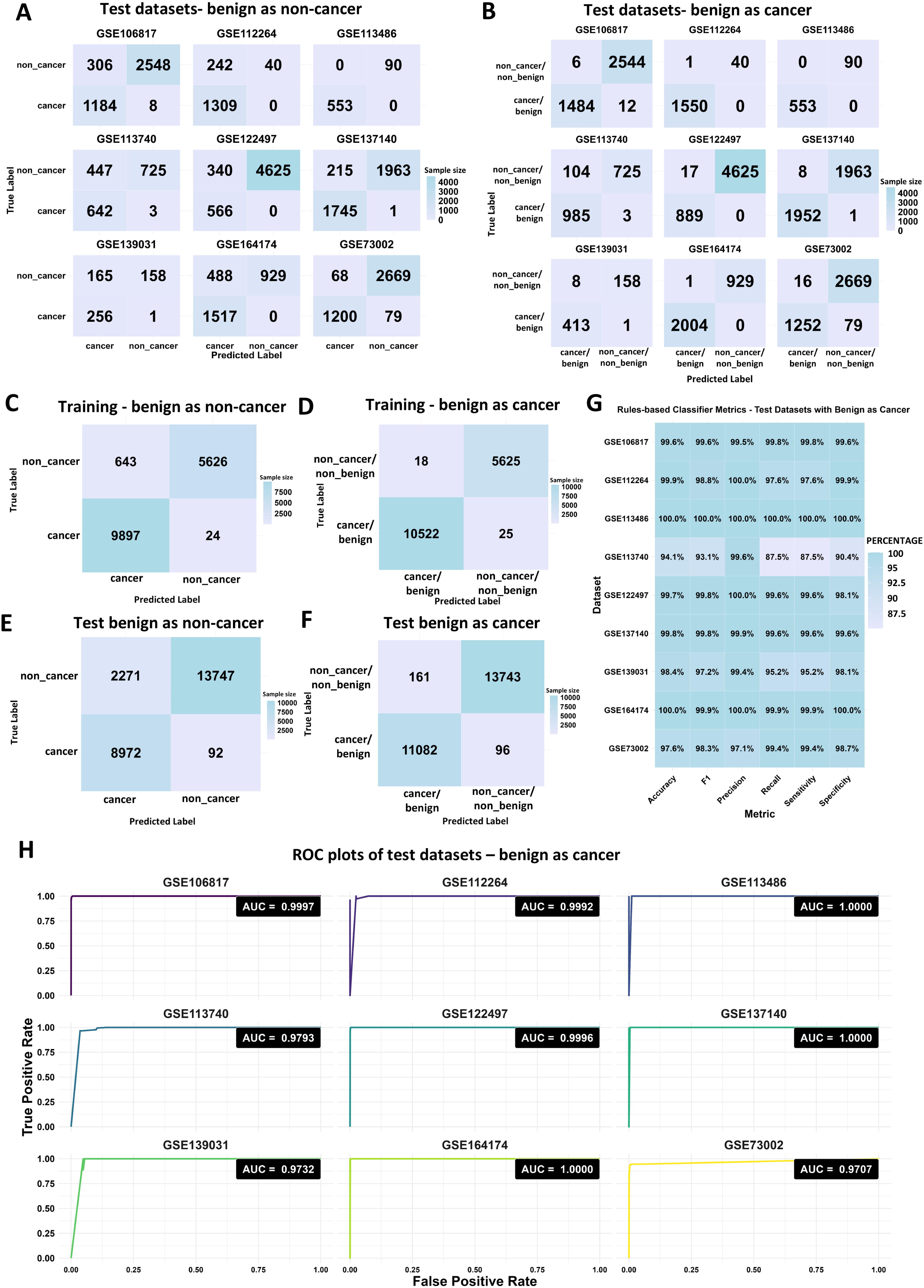
Pan-cancer classifier predicts the cancer and pre-cancerous benign conditions with highest accuracy: The confusion matrices of individual test datasets where (A) benign cases are considered “non_cancer” and (B) “cancer/benign” classes were depicted. The confusion matrices of training data when (C) benign cases are considered “non_cancer” and (D) “cancer/benign” classes, test datasets (all samples from each datasets) as (E) “non_cancer” and (F) “cancer/benign” depicts the improved classification of “cancer/benign” class. (G) The classifier performance metrics when benign cases are considered “cancer/benign” class is depicted as heatmap. (H) The ROC plots depict the highly accurate diagnostic performance in differentiating “cancer/benign” and “cancer/non-benign” classes.

The performance metrices of the test datasets when benign was categorized as cancer/benign plotted through the heatmap suggest the best performance of the pan-cancer classifier (**Fig. 6G**). The overall performance increased very significantly and the ROC plots denotes the excellent diagnostic potential of the classifier at single-sample level (**Fig. 6H**).

Taken together, the rules-based single-sample pan-cancer serum classifier developed in this study achieved higher performance over a massive sample size of 46349 samples from 18 independent studies. Although many transcriptome-based classifier models were identified in several studies, no study has identified pan-cancer serum-based classifier that were evaluated in such a larger sample size. The uniqueness of the single-sample pan-cancer classifier model identified in this study is that the classifier can predict the presence of cancer as well as pre-cancerous benign lesions containing neoplastic cells sample-wise and hence can be intervened for screening programmes for further biochemical characterization of the suspected individuals with least invasive compared to the current available screening and diagnosis methods.

## Discussion

Liquid biopsy allows a minimally invasive way of early diagnosis of cancer and thus can help in reduction of cancer-associated mortality. The existing methods of cancer detection involves the utilization of highly-invasive methods that are expensive and hence not suitable for large scale screening for cancer. The most important drawback of the existing liquid biopsy diagnosis methods that utilizes the cancer-associated mutations is the prevalence of false-positive results^27^. Cancer-associated databases such as TCGA has only the data derived from various cancer tissues and miRNA expression resource is needed for liquid biopsy in various cancer types. This study performed data mining and curation to build a web resource that have expression of miRNAs in 46349 clinical serum samples. This resource will be valuable for any other studies that need to estimate the expression of miRNAs in cancer and other disease conditions in the serum of clinical subjects.

Next, we proposed to identify the miRNA-based classifier that can accurately discriminate between “cancer” and “non_cancer” samples. There are limited or no studies that explored the possibility of a pan-cancer classifier that can identify the presence of cancer directly from serum. The existing studies utilizing machine learning approach to identify the miRNA-based classifier to discriminate between “cancer” and “non_cancer” classes have focused mostly on the correct prediction of cancer. It is equally important to correctly identify the other disease samples as “non_cancer” as the same miRNAs can be dysregulated in cancer as well as other pathological conditions. The classifier model evaluation is limited by the availability of suitable datasets with both the “cancer” and “non_cancer” class comprising other disease conditions. The conventional ML classifier performances are affected by the class imbalance, normalization methods as the ML algorithms identify the “cancer” and “non_cancer” class-specific discriminatory features. To overcome the limitations of the conventional ML classifiers, this study utilized the approach of single-samples rules-based classification algorithms that relies on the relationship of miRNA pairs within the samples and predict the “cancer” or “non_cancer” class. The serum miRNA resource built in this study were utilized for the training of the classifier model and evaluation in test and validation datasets consisting of independent datasets comprising the samples belonging to different cancer types, non-cancer samples including several other diseases.

A total of eight miRNAs constituting 4 miRNA pairs were identified to be accurately discriminating between the “cancer” and “non_cancer” class. The voting-based approach was utilized for the prediction of the class in which more than 2 rules satisfying represents the “cancer” class, otherwise, “non_cancer” class. miR-4258 was observed to have very high expression only in serum but not in whole blood, denoting extra-blood origin in an individual study^28^. miR-4258 expression was observed to be upregulated in various cancers and play an oncogenic role by contributing to metastasis in liver cancer^29,30^. miR-4730 was observed to play a tumor suppressive role in liver cancer ^31^ and the lesser expression is associated with poorer survival in pancreatic cancer ^32^. In our study, we identified the expression of oncogenic miR-4258 was observed to be higher than the expression of tumor-suppressive miR-4730 in “cancer” samples whereas the reverse trend was observed in “non_cancer” samples. It is speculated that the pathology associated with the cancer might lead to the induction of miR-4730 and reduction of miR-4730 and this change can reflect the tumor status. The second rule was miR663a > miR-1228-5p to be predicted as cancer. miR-663a was found to play a tumor promoting role in nasopharyngeal carcinoma and its upregulation was observed to be associated with poorer prognosis. Interestingly, miR-663a is observed to play a tissue specific role in cancer as it promotes malignancy in prostate cancer while it suppresses in pancreatic cancer. It plays a major role related to inflammation genes such as TGFB1 ^33^. miR-1228-5p is said to be enriched in exosomes and plays an important role in cell migration and proliferation by interacting with an enzyme DUSP22, which regulates cell proliferation. Down-regulation of DUSP22 was observed in the cells where miR-1228-5p was high in small cell lung cancer in-vivo studies, leading to increase in migration and proliferation of cancerous cells ^34^. The role of miR-6787-5p, miR-6800-5p, miR-1469 and miR-1268a in cancer diagnosis is yet to be explored.

Initially the pan-cancer classifier performance was observed to be affected while predicting certain “non_cancer” samples that were misclassified as “cancer” samples. After exploring the distribution of incorrect predictions, it was observed those samples were majorly of benign nature. This is interesting in the perspective of early diagnosis applications as the screening of pre-cancerous/ benign samples that are associated with initiation of neoplastic growth and may have potential to turn to cancerous at the early stage is crucial for the improved diagnosis before the advent of the tumor proliferation. This classifier can be used as screening of the high-risk subjects who can be recommended for further clinical diagnosis cancer methods.

This study also has several limitations. Since the study is based on the availability of miRNA expression datasets from the clinical samples available in the public databases, it is restricted only to serum due to the increased availability. There are few reports that suggested the miRNA expression changes between serum, plasma and whole blood and hence this classifier cannot be generalized for all the liquid biopsy miRNA expression. The availability of small-RNA sequencing datasets derived from the serum samples were limited and also the expression of the miRNAs was dependent on the depth of sequencing and library preparation. Hence, the future work is planned to design the custom panel for these miRNAs and identify the rules for the prediction of “cancer” and “non_cancer” samples from the serum samples.

## Methods

### Clinical datasets screening, selection and acquisition

The public transcriptome databases such as Gene Expression Omnibus (GEO) and array express were queried with the keyword “serum miRNA” to retrieve the miRNA transcriptome datasets derived from the serum of the clinical subjects. This search resulted in 68,022 samples across 1495 datasets. The exclusion criteria include the samples that are derived from the cell-line models, non-serum tissue origin including those from exosomes and single-cell RNA-seq datasets. Only the datasets with minimal sample size of 500 was selected for train and test datasets. Duplicates or samples with insufficient metadata information were removed from the analysis. A total of 46,349 samples across 18 independent studies were further selected for the study (**Supp. Fig. 1**) (**Supp. Table 1**).

### Study design

The dataset GSE211692 was utilized as training dataset due to its diverse representation of 13 different cancer types and an adequate sample size for each cancer type, along with the non-cancer controls. The test datasets were independent datasets that comprises both the “cancer” and “non_cancer” classes within each dataset to evaluate the performance of the trained model. The validation datasets comprise of the samples that are having either “cancer” or “non_cancer” class of different disease types. The performance of the classifier in the identification of other disease classes as “non_cancer” class can be explored through the validation datasets.

### Training dataset

GSE211692 (n = 16,190)

### Test datasets

GSE122497, GSE106817, GSE137140, GSE113740, GSE112264, GSE139031, GSE113486, GSE164714, and GSE73002 (n = 25,508)

### Validation datasets

GSE110317, GSE110651, GSE134108, GSE85589, GSE117064, GSE120584, GSE150693, and GSE140249 (n = 5,077)

The datasets included a variety of cancer types including, benign conditions, and non-cancer diseases such as Autoimmune Hepatitis, Alzheimer’s disease (AD), Vascular Dementia (VD), and Mild Cognitive Impairment (MCI). (**Supp. Table 1**).

### Dataset retrieval and pre-processing

The miRNA expression matrices and the metadata information about the clinical subjects were retrieved through the R package GEOquery^35^. In training data, only the miRNAs that are expressed in at least 10% of samples were retained. This is done to ensure that the expression of miRNAs should be at higher level in serum and hence after the training of the classifier model, the expression the miRNAs must be there in test and validation datasets too. A total of 211 miRNAs with higher expression was remained for the further analysis. Batch effect correction was not required in the case of test datasets as the classifier was evaluated as single sample level rather than the dataset level.

### Training of the pan-cancer classifier

The rules-based single-sample classifier was developed based on the pairwise comparisons between the miRNAs. The top scoring pairs (TSPs) for the samples belonging to the “cancer” and “non_cancer” class were identified. If there are “N” number of miRNAs in the expression matrix, the total number of possible pairs per class for the first miRNA will be N-1. The total possible number of pairs per class will be N(N-1)/2 pairs. For “m” number of samples, the total number of possible pairs per class will be m*N(N-1)/2 pairs. There are a total 16190 samples in the training dataset and the number of highly expressed miRNAs after the filtration step was 211 miRNAs. So, for each class there will be 211*210/2 = 22155 pairs was calculated. For “cancer” and “non_cancer” the combined restricted pairs will be 22155+22155 = 44310 pairs. For 16190 samples, a total of 3.5*10^8 pairs were calculated for “cancer” class and 3.5*10^8 pairs were calculated for “non_cancer” class leading to the analysis of around 7.17*10^8 pairs. The top scoring pairs for each class was calculated by the R packages, switchBox^36^ and multiclassPairs^37^.

A total of 4 rules were identified based on the scores that gives the idea about the trajectory specific for the particular class. The classification rules were determined by the miRNA expression relationships:

For example, if the expression of miRNA 4258 > miRNA 4730, then the sample will be classified as “cancer”. If the expression of miRNA 4730 > miRNA 4258, the sample will be classified as “non_cancer”. The miRNA names are mentioned only with the number and 5p or 3p if applicable for the sake of better annotation during the training.

A total of 4 TSPs were identified and a voting-based approach was implemented to identify the class of the sample with higher accuracy. If more than two rules out of 4 rules for “cancer” class were satisfied, then it will be classified as “cancer”. If not, it will be classified as “non_cancer”.

### Performance Evaluation

The Evaluation of the performance of the pan-cancer serum classifier was performed by calculating the performance metrices such as Sensitivity, specificity, accuracy, Precision and F1-score. The confusion matrices were plotted to depict the relationship between the correct predictions of “cancer” class – True-positive (TP) – “cancer” identified as “cancer”, True-negative (TN) – “non_cancer” predicted as “non_cancer” and the incorrect predictions – False-postive (FP) – “non_cancer” predicted as “cancer”, False-negative (FN) – “cancer” predicted as “non_cancer”.

Sensitivity measures the classifier ability to predict the “cancer” samples correctly. Higher sensitivity denotes the lesser “cancer” samples are misclassified as “non_cancer”.

### Sensitivity = TP/(TP+FN)

Specificity measures the ability of the classifier to identify the non-cancer samples correctly as “non_cancer”. A high specificity denotes lesser “non_cancer” samples misclassified as “cancer”.

### Specificity = TN/(TN+FP)

Accuracy measures the proportion of samples predicted correctly out of all the samples tested. The higher value of accuracy represents the overall correctness of the classifier.

### Accuracy = (TP+TN)/(TP+TN+FP+FN)

Precision measures how many of the predicted cancer cases were actually cancer. Higher precision means fewer “non_cancer” samples were misclassified as cancer.

### Precision = TP/(TP+FP)

Recall measures the ability of the classifier to correctly identify the positive instances. Higher recall value depicts the lesser false negatives.

### Recall = TP/(TP+FN)

F1-score measures the balance between the sensitivity and precision. It is important when the dataset is affected by class imbalance.

### F1-score = 2 * [(Precision * Sensitivity)/ (Precision + Sensitivity)]

The confusion matrices were plotted in form of heatmap representing the total number of samples and all the metrices were calculated and plotted using the Machine learning R package caret. For the better visualization all the metrices are given as percentages in the plot.

### ROC analysis

Receiver Operating characteristics (ROC) curve plots were plotted to represent the relation between the true positive rate (y-axis) and the false positive rate (x-axis). ROC analysis was performed and visualized through the R package pROC^38^.

The classifier’s robustness was assessed across independent test and validation datasets. The impact of benign samples on classification performance was analyzed by reassigning the “cancer” class to “cancer/benign” class by including the benign samples and “non_cancer” class to “non_cancer/non_benign” by excluding benign samples.

### Visualization methods

For the distribution of samples in each dataset, t-SNE plots were plotted through the R package “Rtsne”. For better understanding all the “cancer” class was represented in red color and “non_cancer” class was represented in blue color. The distribution plots for the training, test and validation datasets were plotted as lollipop and tile plots that tells the relationship between the number of samples in each disease state. The expression of eight miRNAs forming the four rules were visualized using beeswarm plots plotted through the R package “ggbeeswarm”. Bee-swarm plots used for the better visualization in distinguishing individual data points and prevent over-plotting, especially in the datasets with larger sample sizes. The incorrect predictions across the training, test and validation datasets were plotted as bar plots denoting the number of correct and wrong predictions in each dataset. The R packages ggplot2, dplyr, tidyverse were used for the plots and data-frame operations.

All the analysis was performed using R 4.4.1 version in a computational workstation with I9 processor with 32 threads and 128GB RAM physical memory.

### Data and code availability

All the datasets used in the study are publicly available from the GEO database. The codes used for the analyses performed in the study can be requested upon the contacting the corresponding author.

## Data Availability

All data produced in the present work are contained in the manuscript and further requests can be available upon reasonable request to the authors.

## Acknowledgement

This work was supported by the intramural research grant from IISER Bhopal and the support for the computational workstation from Indian Institute of Science Education and Research, Bhopal (IISER Bhopal). We thank all the authors who have deposited the miRNA transcriptome datasets in public databases.

## Author information

**Authors and Affiliations:**

**Indian Institute of Science Education and Research, Bhopal (IISER Bhopal)**

Pandikannan Krishnamoorthy

Madhavan Parthasarathy

Nilanjana Das

Himanshu Kumar

**All India Institute of Medical Sciences, Bhopal (AIIMS Bhopal)**

Ashok Kumar

Vikas Gupta

Saikat Das

**Frontiers Research Center, Osaka University**

Himanshu Kumar

## Contributions

P.K and H.K. conceived the project. P.K, M.P, N.D and ASR contributed in the datasets screening. P.K. optimized and formulated the methodology and wrote the codes for the analysis. P.K and M.P. did the formal analysis and visualization. M.P and P.K annotated and developed the web resource. P.K, M.P, N.D and H.K wrote the original draft of the paper. A.S, V.G and S.D contributed to the review of the study design, P.K, M.P, N.D, ASR and H.K reviewed and edited the paper. H.K. supervised the entire project. All authors read and approved the final paper.

## Ethics declarations

All the analyses are made from the publicly available clinical transcriptome datasets. No patients were recruited nor any experiment is performed on clinical subjects and hence requirement of Institute Ethical committee is not required for this study.

H.K and P.K has a patent registered related to this work.

## Supplementary Figures Legend

**Supp. Fig. 1:**

**Serum samples and selection criteria:** Serum miRNA datasets sample-wise selection strategy and criteria following PRISMA guidelines.

**Supp. Fig. 2:**

**Sample distribution in test datasets:** t-distributed Stochastic Neighbour Embedding (t-SNE) plots depicting the distribution of samples in test datasets between cancer and non_cancer classs in (A) GSE106817, (B) GSE112264, (C) GSE113486, (D) GSE113740, (E) GSE122497, (F) GSE137140, (G) GSE139031, (H) GSE164174, (I) GSE73002.

**Supp. Fig. 3:**

**Expression of classifier miRNAs in test datasets:** Expression of 8 miRNAs constituting the classifier rules in test datasets (A) GSE106817, (B) GSE112264, (C) GSE113486, (D) GSE113740, (E) GSE122497, (F) GSE137140, (G) GSE139031, (H) GSE164174, (I) GSE73002. Red denotes the cancer class while the blue denotes the non_cancer class. y-axis denotes the normalized expression values and x-axis denotes the class names.

**Supp. Fig. 4:**

**Classification of true and predicted cancer and non_cancer in test datasets:** Confusion matrices depicting the comparison between the classifier results and the actual labels as explained in (A), for the 4 rules individually in all the 9 test datasets (B-J).

**Supp. Fig. 5:**

**Classification of true and predicted cancer and non_cancer in test datasets:** Confusion matrices depicting the comparison between the classifier results and the actual labels as explained in (A), for the 4 rules individually in all the 8 Validation datasets (B-I). (J) The heatmap depicts the prediction results of all the samples that are part of validation datasets comprising different disease types.

**Supp. Fig. 6:**

**Benign Sample distribution in test datasets:** t-distributed Stochastic Neighbour Embedding (t-SNE) plots depicting the distribution of samples in test datasets between “cancer/benign” and “non_cancer” classes in (A) GSE106817, (B) GSE112264, (C) GSE113486, (D) GSE113740, (E) GSE122497, (F) GSE137140, (G) GSE139031, (H) GSE164174, (I) GSE73002. The benign samples form a distinct distribution.

**Supp. Fig. 7:**

**Classification of true and predicted cancer, benign, and non_cancer in test datasets:**Confusion matrices depicting the comparison between the classifier results and the actual labels between cancer/benign as one class and non_cancer/non_benign as another class as explained in (A), for the 4 rules individually in all the 9 test datasets (B-J).

**Supp. Fig. 8:**

**Model Performance after considering benign and pre-cancerous lesion as cancer:** The classifier performance metrics such as Specificity, Sensitivity, Recall, Precision, F1 and Accuracy are plotted as heatmap. The comparison between the performance when benign is considered as non_cancer (A) and when benign is considered as cancer (B) is depicted.

## Supplementary Tables

**Supp. Table 1:**

The training, test and validation datasets used in the study.

**Supp. Table 2:**

Prediction results of training dataset by the serum pan-cancer classifier.

**Supp. Table 3:**

Prediction results of test datasets by the serum pan-cancer classifier.

**Supp. Table 4:**

Prediction results of validation datasets by the serum pan-cancer classifier.

**Supp. Table 5:**

Prediction results of training dataset when benign is categorized in cancer/benign class by the pan-cancer classifier.

**Supp. Table 6:**

Prediction results of test datasets when benign is categorized in cancer/benign class by the pan-cancer classifier.

