## Supplementary figures and images for "Global Pan-cancer serum miRNA classifier across 13 cancer types: Analysis of 46,349 clinical samples"

### Supp. Fig. 1

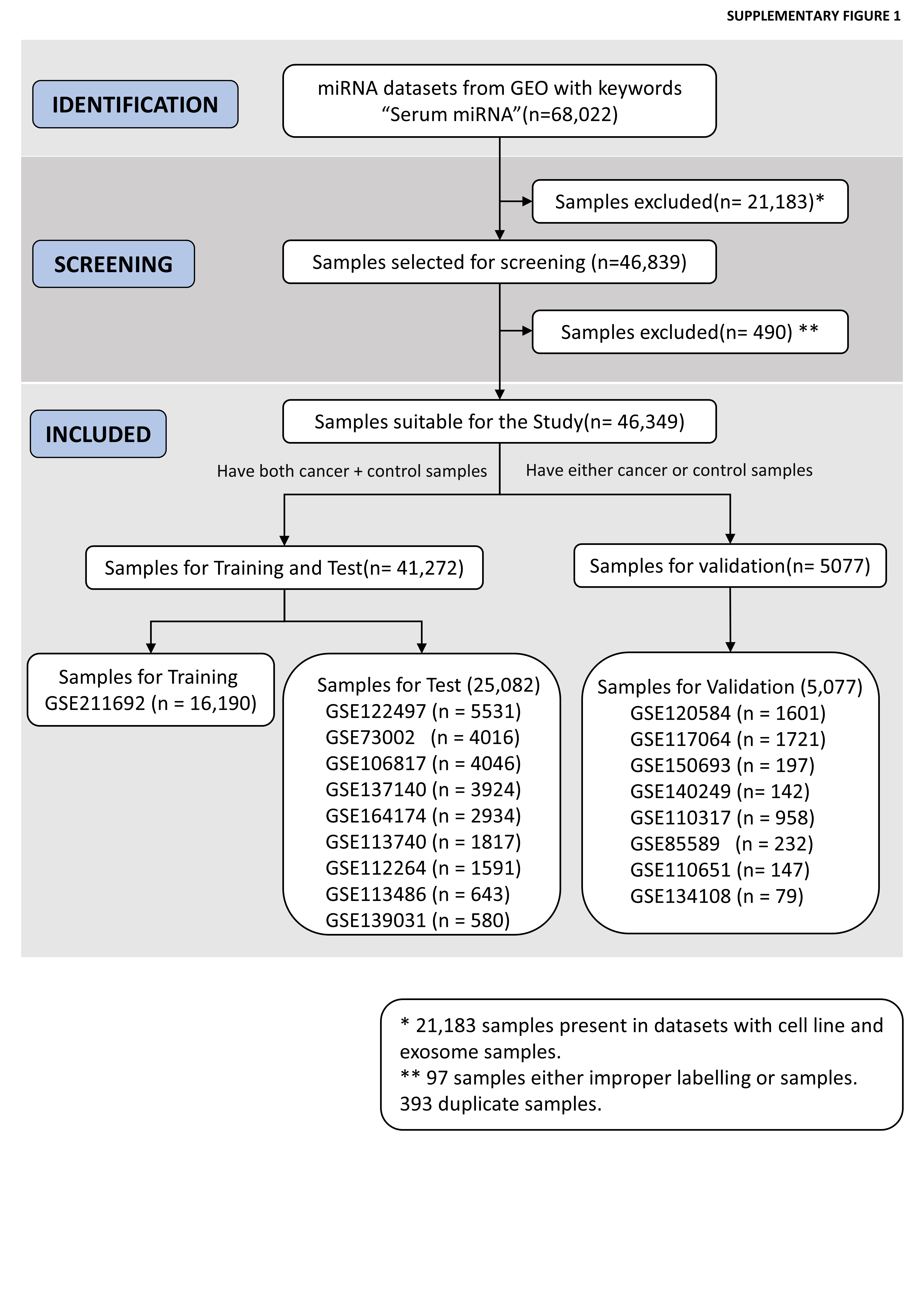

### Supp. Fig. 2

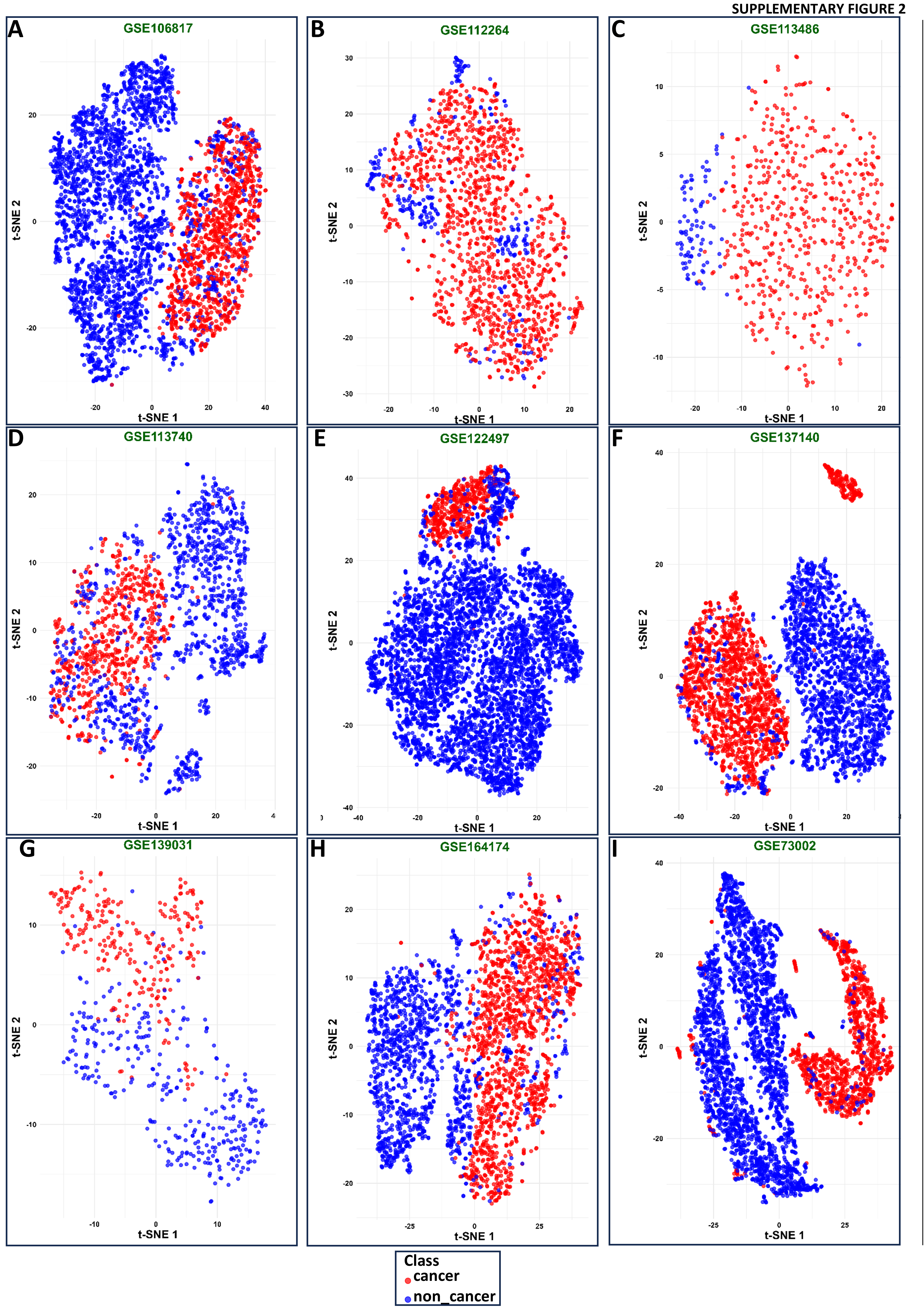

### Supp. Fig. 3

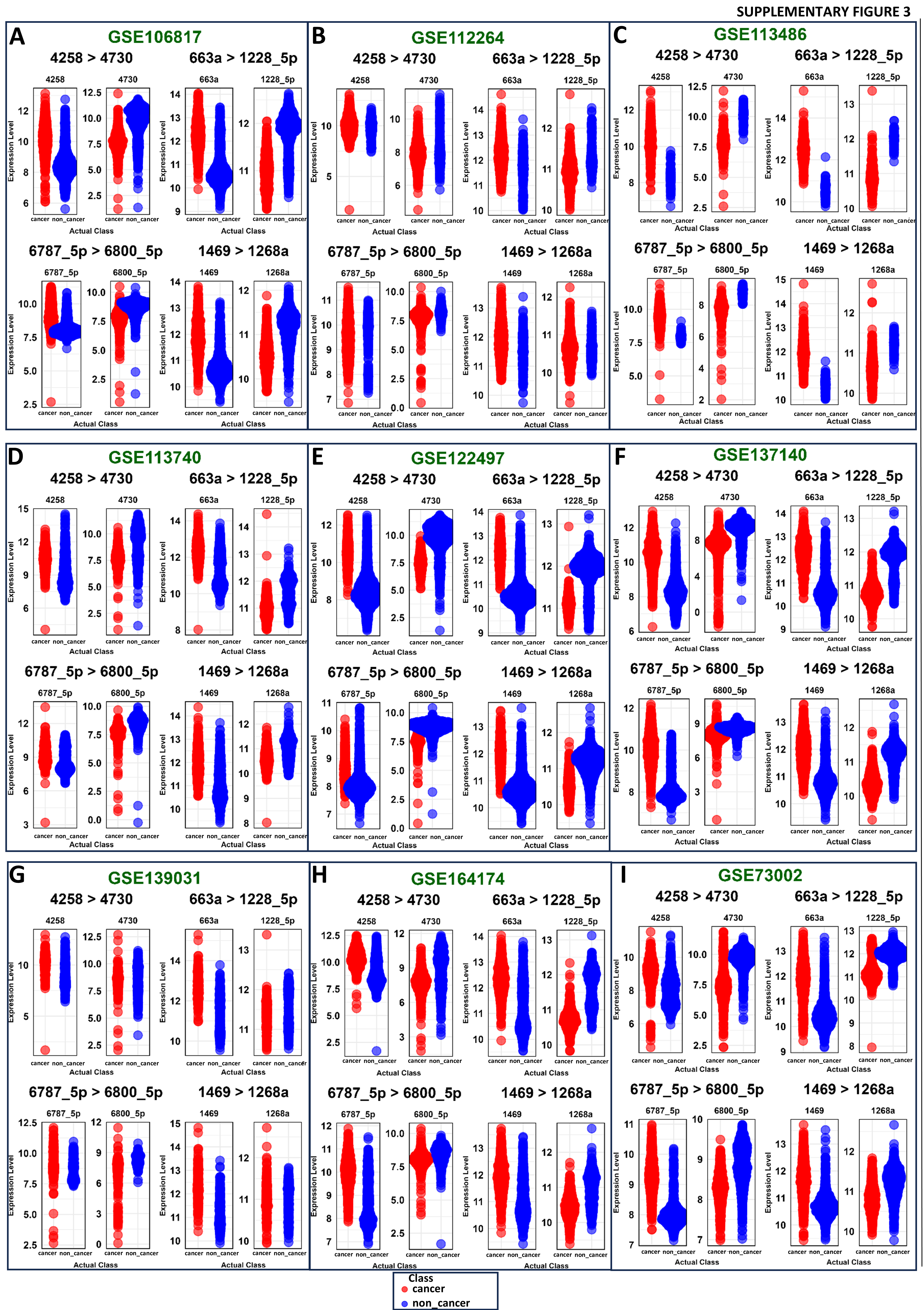

### Supp. Fig. 4

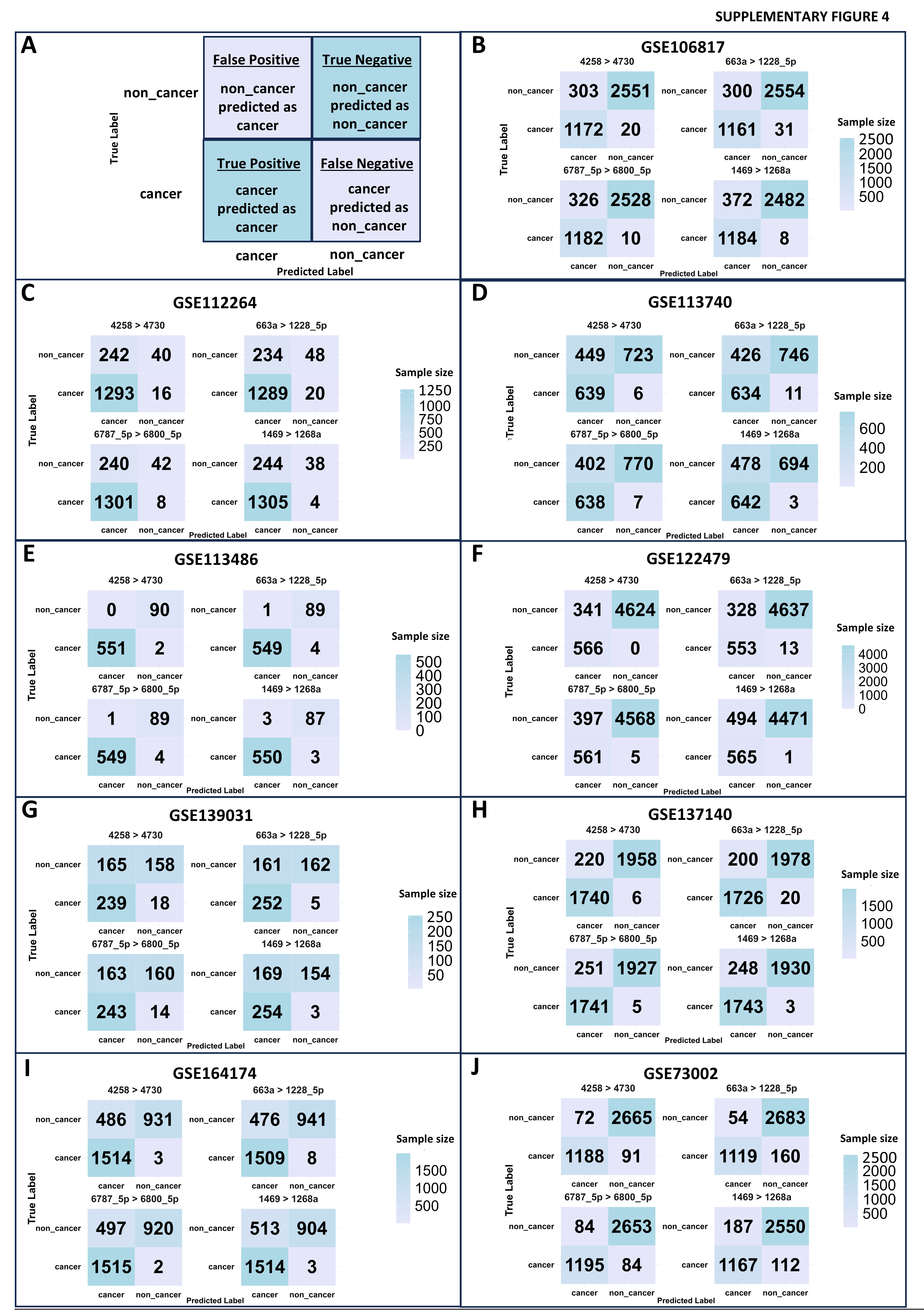

### Supp. Fig. 5

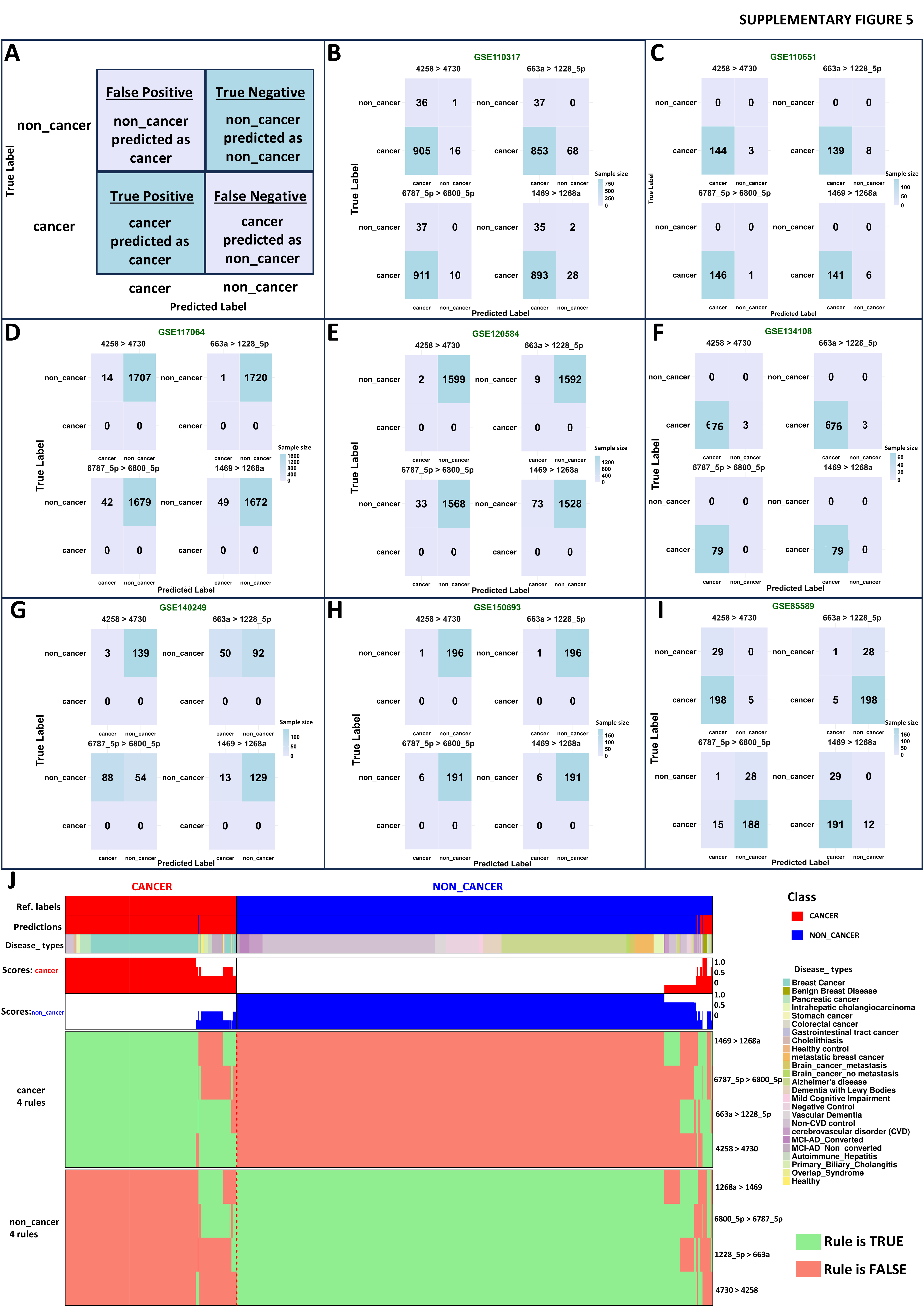

### Supp. Fig. 6

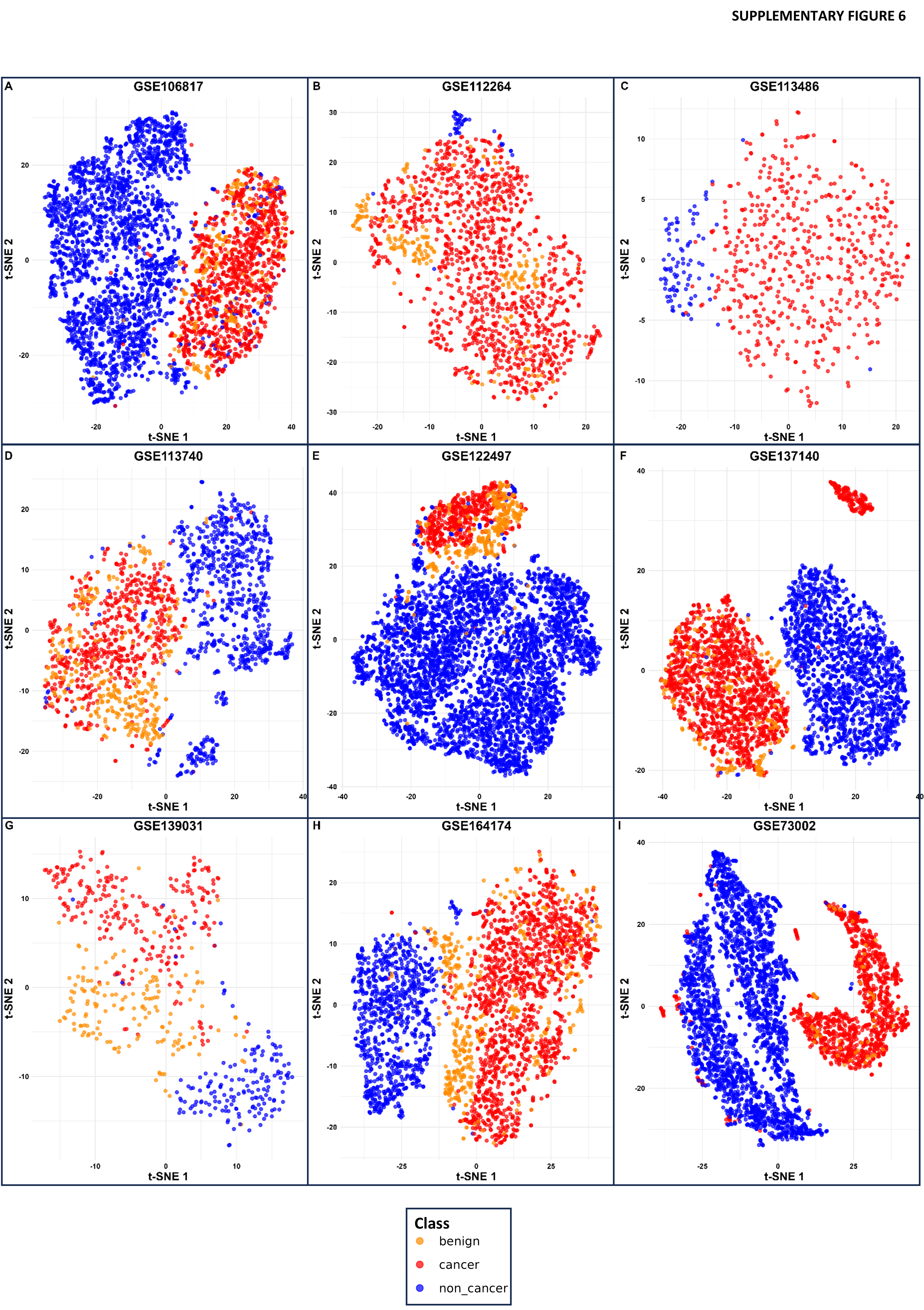

### Supp. Fig. 7

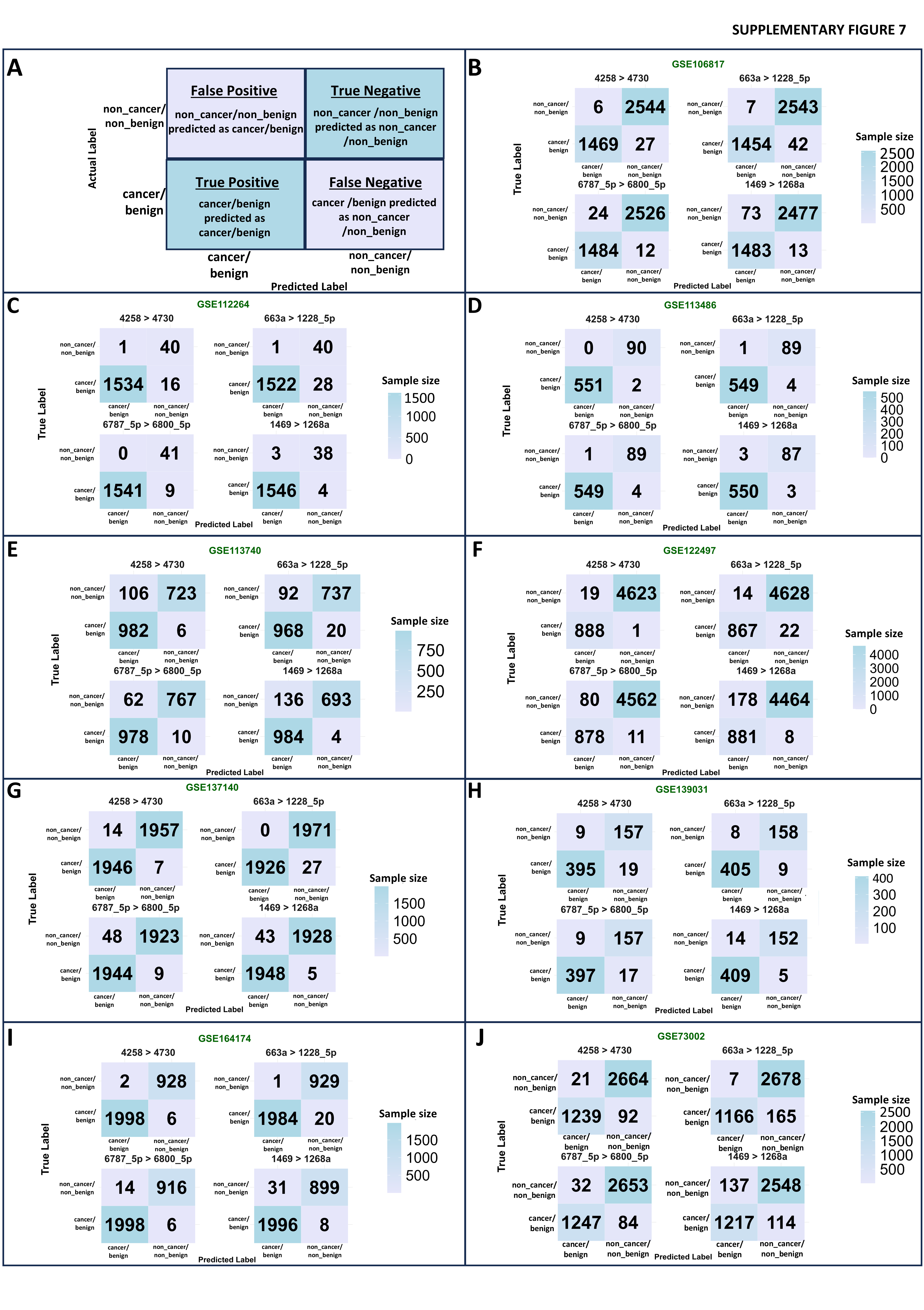

### Supp. Fig. 8

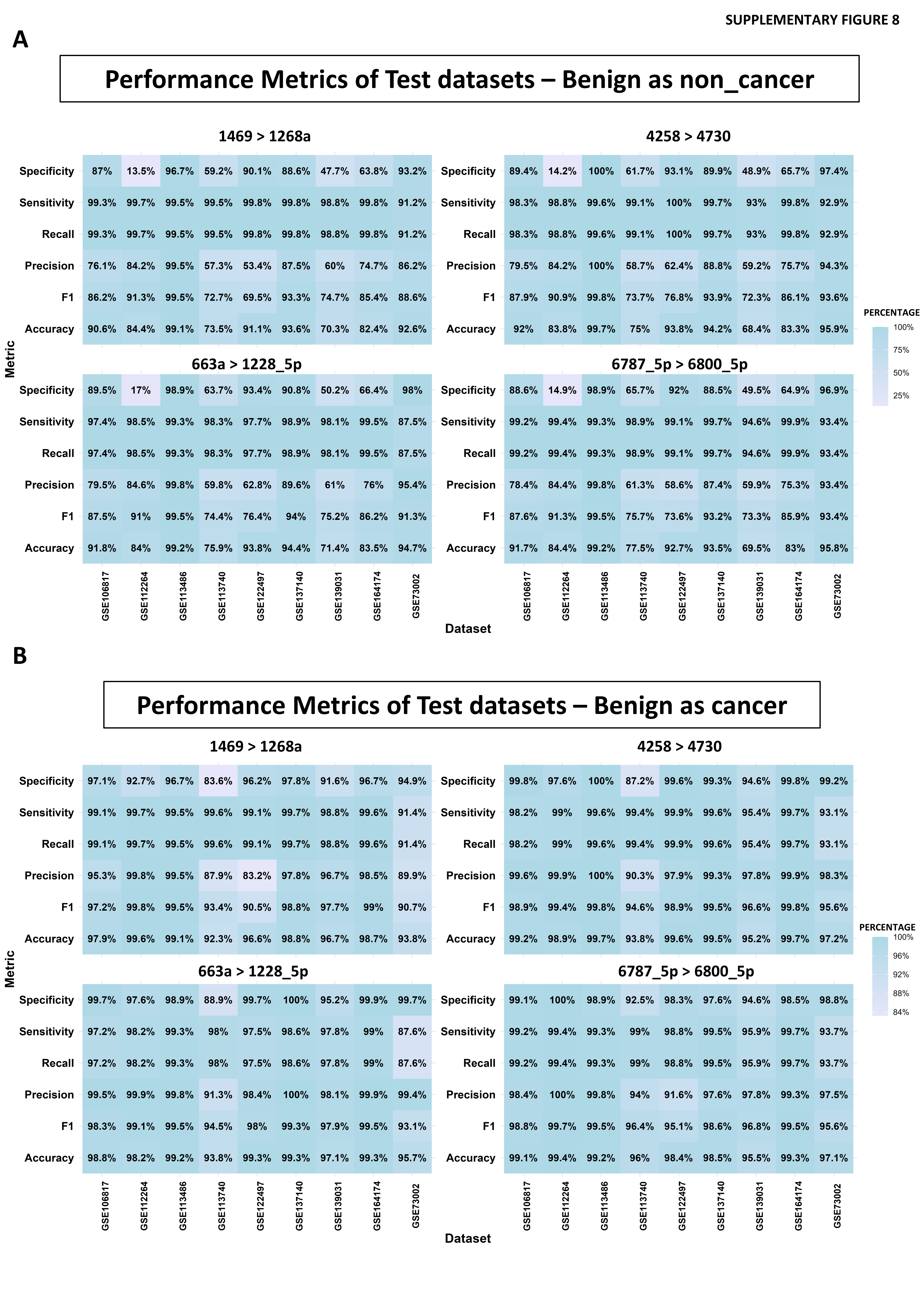
